# Sex and Education Modify the Association Between Subjective Cognitive Decline and Amyloid Pathology

**DOI:** 10.1101/2023.11.03.23297795

**Authors:** Corey J. Bolton, Omair A. Khan, Dandan Liu, Sydney Wilhoite, Logan Dumitrescu, Amalia Peterson, Kaj Blennow, Henrik Zetterberg, Timothy J. Hohman, Angela L. Jefferson, Katherine A. Gifford

## Abstract

**Background:** Subjective cognitive decline (SCD) may be an early risk factor for dementia, particularly in highly educated individuals and women. This study examined the effect of education and sex on the association between SCD and Alzheimer’s disease (AD) biomarkers in non-demented older adults.

**Method:** Vanderbilt Memory and Aging Project participants free of clinical dementia or stroke (n=156, 72±6 years, 37% mild cognitive impairment, 33% female) completed fasting lumbar puncture, SCD assessment, and Wide Range Achievement Test-III Reading subtest to assess reading level at baseline as a a proxy for educational quality. Cerebrospinal fluid (CSF) biomarkers for AD (β-amyloid_42_ (Aβ42), Aβ42/40 ratio, phosphorylated tau (p-tau), tau, and neurofilament light (NfL)) were analyzed in batch. Linear mixed effects models related SCD to CSF AD biomarkers and follow-up models assessed *SCD x sex, SCD x reading level*, and *SCD x education* interactions on AD biomarkers.

**Result:** In main effect models, higher SCD was associated with lower Aβ42 and Aβ42/40 ratio (p-values<0.004). SCD was not associated with tau, p-tau, or NfL levels (*p-*values>0.38). SCD score interacted with sex on Aβ42/40 ratio (*p*=0.03) but no other biomarkers (*p*-values>0.10). In stratified models, higher SCD was associated with lower Aβ42/40 ratio in men (*p*=0.0003) but not in women (*p*=0.48). SCD score interacted with education on Aβ42 (*p*=0.005) and Aβ42/40 ratio (*p*=0.001) such that higher education was associated with a stronger negative association between SCD and amyloid levels. No *SCD score x reading level* interaction was found (p-values> 0.51) though significant associations between SCD and amyloid markers were seen in the higher reading level group (p-values<0.004) but not the lower reading level group (p-values>0.12) when stratified by a median split in reading level.

**Conclusion:** Among community-dwelling older adults free of clinical dementia, higher SCD was associated with greater cerebral amyloid accumulation, one of the earliest pathological AD changes. SCD appears most useful in detecting early AD-related brain changes in men and individuals with higher quantity and quality of education. SCD was not associated with CSF markers of tau pathology or neurodegeneration. These findings suggest that considering sex and education is important when assessing SCD in older adults.

## Introduction

Alzheimer’s disease (AD) is a public health crisis that will only continue to worsen as the population ages.^1^ Novel treatments for AD require initiation prior to the onset of dementia, highlighting the need for early detection in individuals who are cognitively unimpaired or only mildly symptomatic.^2^ Current methods of identifying underlying AD pathology in non-demented individuals, such as positron emission tomography imaging and lumbar puncture, can be expensive and would place an unsustainable burden on the healthcare system if implemented widely. Screening patients to identify those at elevated risk is one way to dramatically reduce the cost associated with early identification of AD pathology.^3^ Screening measures that are efficient and cost-effective are essential to sustain the current needs for early identification in AD.

One efficient and inexpensive method of screening and identifying patients at increased risk of AD is through utilizing measures of subjective cognitive decline (SCD). Higher levels of SCD have been associated with cognitive decline,^4,5^ hippocampal atrophy,^6^ changes in cerebral blood flow,^6^ and progression to MCI and dementia.^7^ Despite the utility of SCD in predicting AD-related brain and clinical changes, there are numerous factors beyond AD that can contribute to SCD and many clinical and demographic factors that may modify the ability of SCD to predict AD-related changes. For example, SCD has been more strongly associated with clinical decline in women than in men,^8^ and in individuals with higher levels of education compared to those with lower levels of education.^7^ SCD has also been associated with cerebrospinal fluid (CSF) biomarkers of AD;^9^ however, this association is inconsistent,^10^ possibly due to demographic differences. Despite recognition that clinical and demographic factors may influence AD biology and clinical manifestation, there is a paucity of work examining the specific factors and nature of the effect modification of these factors on the ability of SCD to predict underlying AD pathology in non-demented patients.

This study seeks to examine the associations between a novel SCD measure and CSF biomarkers of AD in non-demented older adults, and to determine the effect of common demographic factors on this association. Based on past work,^7–9^ we are focusing on sex and education as potentially modifying factors. We hypothesize that this novel SCD measure will be associated with CSF biomarkers of AD and that these associations will be stronger in women. We will also investigate two different markers of education, including years of education completed and a single-word reading metric as a proxy for educational quality. We hypothesize SCD and CSF biomarker associations will be stronger in individuals with more years of education and greater educational quality. We aim to identify which factors influence the association of SCD and underlying AD pathology to aid in identifying patients in whom novel interventions may be most beneficial.

## Methods

### Cohort

Participants were drawn from the baseline cohort of the Vanderbilt Memory and Aging Project, a longitudinal study investigating vascular health and brain health among aging adults.^11^ Inclusion criteria required participants to be age 60 or older, speak English, have adequate visual and auditory acuity, and have a reliable study partner. To determine study eligibility, participants completed a medical history review, clinical interview, and neuropsychological assessment. Cognitive diagnosis was determined by consensus, including normal cognition (NC), early mild cognitive impairment (eMCI),^12^ or MCI based on the National Institute on Aging/Alzheimer’s Association Workgroup clinical criteria.^13^ Participants were excluded for magnetic resonance imaging (MRI) contraindication, history of neurological disease (e.g., dementia, stroke), major psychiatric illness, heart failure, severe head injury (loss of consciousness ≥5 minutes), and systemic or terminal illness (e.g., cancer) that could affect follow-up participation. At study enrollment, participants completed a comprehensive evaluation, including but not limited to physical and frailty examination, fasting blood draw, clinical interview, SCD module, echocardiogram, brain MRI, and optional lumbar puncture. Participants were excluded from the current analyses for missing baseline SCD, covariate, or CSF data.

The protocol was approved by the Vanderbilt University Medical Center Institutional Review Board, and written informed consent was obtained from all participants prior to data collection. Due to participant consent limitations in data sharing, a subset of data is available for purposes of reproducing the results or procedures. These data, analytic methods, and study materials can be obtained by contacting the corresponding author.

### SCD Questionnaire

Participants completed four questionnaires assessing SCD: the Everyday Cognition Questionnaire,^14^ the Memory Functioning Questionnaire,^15^ the Cognitive Difficulties Scale,^16^ and the Cognitive Changes Questionnaire.^17^ Items from these questionnaires were reduced into a 45-item questionnaire (the Vanderbilt SCD Questionnaire) using psychometric methods including item response theory and computerized adaptive testing.^18^ Scores on this measure range from 38 to 192, with higher scores indicating more SCD. The current study considered SCD score as a continuous measure, as opposed to a dichotomized diagnostic status. This measure was not used to determine study eligibility or cognitive status.

### Lumbar Puncture and Biochemical Analyses

Participants completed an optional fasting lumbar puncture at study enrollment. CSF was collected with polypropylene syringes using a Sprotte 25-gauge spinal needle in an intervertebral lumbar space. Samples were immediately mixed and centrifuged, and supernatants were aliquoted in 0.5mL polypropylene tubes and stored at −80°C. Samples were analyzed in batch using commercially available enzyme-linked immunosorbent assays (Fujirebio, Ghent, Belgium) to determine the levels of amyloid-β_1-42_ (Aβ42; INNOTEST® β-AMYLOID_(1-42)_), Aβ_42_ and Aβ_40_ to calculate the Aβ42/40 ratio (Aβ Triplex Assay, Meso Scale Discovery), phosphorylated tau (p-tau; INNOTEST® PHOSPHO-TAU_(181P)_), and total tau (t-tau; INNOTEST® hTAU). P-tau was measured by tagging a tau phosphorylation site at threonine 181. Neurofilament light (NfL) was measured using a commercially available enzyme-linked immunosorbent assay (Uman Diagnostics). Board-certified laboratory technicians processed data blinded to clinical information, as previously described.^19^ Intra-assay coefficients of variation were <10%.

### Reading Level Assessment

Reading level was assessed at eligibility using the Wide Range Achievement Test 3^rd^ edition (WRAT-III) Reading subtest.^20^ Scores on this measure range from 0 to 57, with 0-41 representing approximately below high school reading level, 42-47 representing high school reading level, and 48-57 representing post-high school reading level. For stratified analyses, reading level was dichotomized by a median split. This test is a commonly used measure to estimate premorbid intelligence and education quality.^21^

### Covariates

The current study adjusted for age, sex, education, race/ethnicity, *APOE-*ε4 status, cognitive status, and score on the Geriatric Depression Scale (GDS).^22^ APOE genotyping was performed using a TaqMan assay on DNA extracted from whole-blood samples,^11^ and APOE-ε4 carrier status was defined as positive (ε2/ε4, ε3/ε4, ε4/ε4) or negative (ε2/ε2, ε2/ε3, ε3/ε3). The following questions related to SCD/cognition were excluded from the GDS score, as these data are likely to confound analyses with SCD as our predictor: “Do you feel you have more problems with your memory than most?”, “Do you have trouble concentrating?”, “Is it easy for you to make decisions?”, and “Is your mind as clear as it used to be?”.

### Analytic Plan

Linear mixed effects models related SCD to CSF AD biomarkers (Aβ42, Aβ42/40 ratio, tau, p-tau, and NfL), adjusting for age, sex, education, race/ethnicity, *APOE-*ε4 status, cognitive status, and GDS score. Follow-up models assessed *SCD x sex*, *SCD x reading level*, and *SCD x education* interactions on AD biomarkers with subsequent models stratified by sex (male, female), reading level split at median (lower half, upper half), and education (lowest tertile, highest tertile), respectively.

Sensitivity analyses excluded predictor or outcome values >4 standard deviations from the group mean to determine if outliers influenced results. Multiple comparison correction was performed across outcomes per model using a false discovery rate based on Benjamini-Hochberg’s procedure. Analyses were performed using R 3.5.2 (www.r-project.org), and significance was set *a priori* at p<0.05.

## Results

### Participant Characteristics

Participants included 129 adults ages 61 to 90 (28% female, 94% non-Hispanic White, 31% *APOE*-ε4 carriers, 16±3 years of education). SCD was significantly correlated with CSF Aβ42 (*r*=-0.30, *p*=0.0006) and Aβ42/40 (*r*=-0.28, *p*=0.001), but not other outcomes (*p*-values>0.06). See **Table 1** for participant characteristics for the entire sample and stratified by sex.

**Table 1.** Participant Characteristics.

|  | Combined (n=129) | Men (n=93) | Women (n=36) | *p-value*<sup>b</sup> |
| --- | --- | --- | --- | --- |
| Age, years | 72.9±7 | 72.6±6 | 73.7±8 | 0.55 |
| Education, years | 16.1±3 | 16.6±3 | 14.6±3 | **<0.001** |
| Race, % non-Hispanic White | 94 | 95 | 92 | 0.53 |
| APOE ε4, % carrier | 31 | 31 | 31 | 0.94 |
| Diagnosis, % MCI | 33 | 30 | 42 | 0.44 |
| GDS score<sup>a</sup> | 2.2±2.6 | 2.1±2.4 | 2.6±3.3 | 0.59 |
| WRAT Reading score | 51.0±5.2 | 51.0±4.6 | 50.8±4.9 | 0.86 |
| SCD score | 62.3±25.3 | 60±21 | 67±27 | 0.38 |
| Aβ42, pg/mL | 553.6±301.4 | 603±279 | 425±204 | **<0.001** |
| Aβ42/40 | 0.89±0.4 | 0.95±0.3 | 0.72±0.3 | **<0.001** |
| Tau, pg/mL | 422.8±223.0 | 403±184 | 473±262 | 0.23 |
| P-tau, pg/mL | 60.7±26.8 | 59±23 | 65±29 | 0.34 |
| NfL, pg/mL | 1098.0±574.4 | 1128±610 | 1018±602 | 0.12 |

Note. Values denoted as mean ± standard deviation or frequency. Bold font indicates *p*-value<0.05. Aβ, amyloid beta; GDS, Geriatric Depression Scale; MCI, mild cognitive impairment; NfL, neurofilament light; p-tau, phosphorylated tau; SCD, subjective cognitive decline; WRAT, Wide Range Achievement Test 3<sup>rd</sup> edition. <sup>a</sup>minus points for cognition. <sup>b</sup>Wilcoxon's test was used for continuous variables, and Pearson's chi-squared test was used for categorical variables.

### SCD and CSF Biomarkers

Greater SCD was associated with lower CSF Aβ42 (β=-3.34, *p*=0.003) and Aβ42/40 (β=-0.004, *p*=0.004) but was not associated with CSF levels of tau, p-tau, and NfL (*p*-values>0.38). These results persisted after FDR correction and were largely unchanged in sensitivity analyses excluding outliers. See **Table 2** for results and **Figure 1** for illustrations.

**Figure 1.**
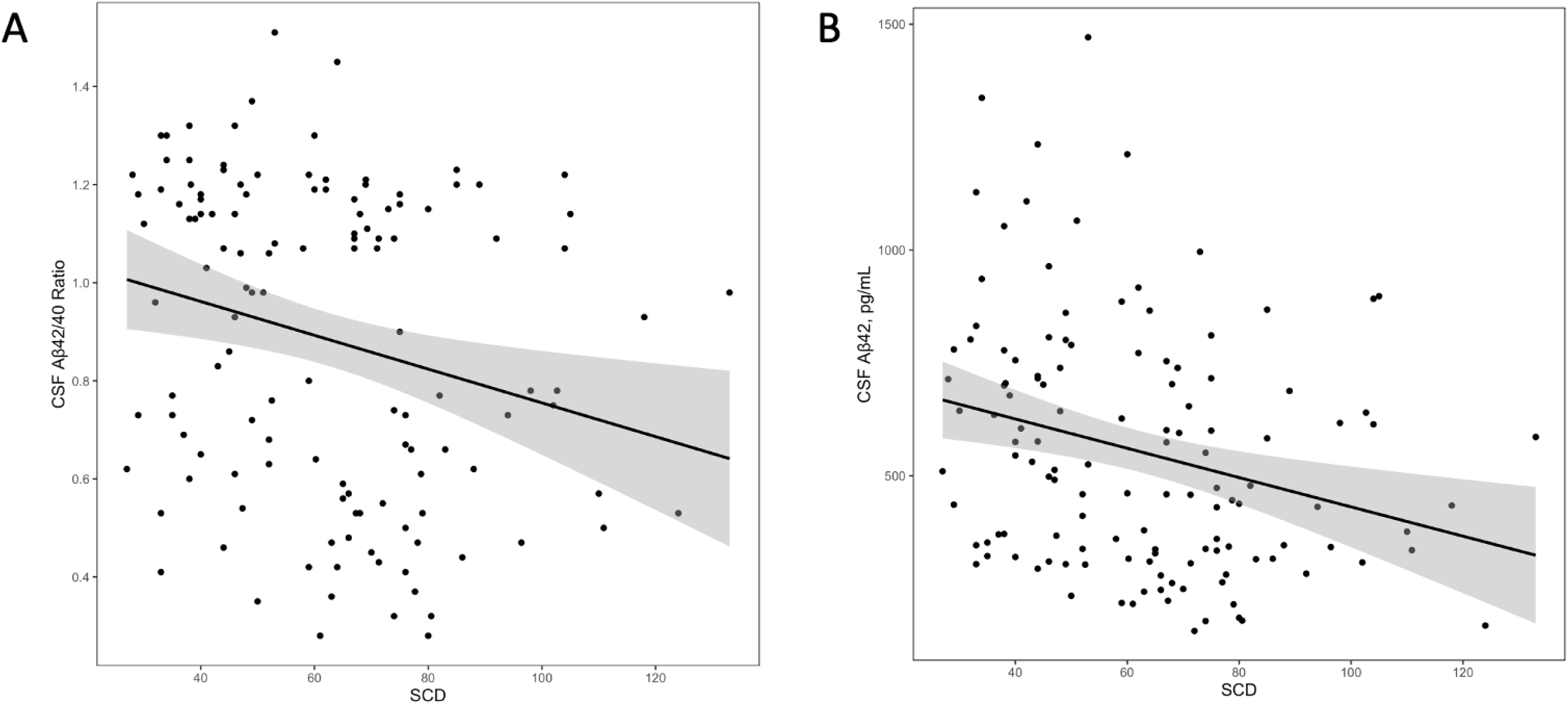
Associations Between Subjective Cognitive Decline and Cerebrospinal Fluid Levels of Amyloid-β_42_ and Amyloid-β_42/40_ Ratio. Lines reflect CSF biomarker values corresponding to SCD levels. Shading reflects 95% confidence interval. (A) Associations between SCD and CSF Aβ42/40 ratio, β=-0.004, p=0.004. (B) Associations between SCD and CSF Aβ42, β=-3.34, p=0.003. Aβ, amyloid beta; SCD, subjective cognitive decline.

**Table 2.** SCD Associations with CSF Biomarkers.

| | $\beta$ | 95% CI | *p* |
| --- | --- | --- | --- |
| Aβ42, pg/mL | -3.34 | -5.54, -1.15 | **0.003\*** |
| Aβ42/40 | -0.004 | -0.006, -0.001 | **0.004\*** |
| Tau, pg/mL | 0.84 | -1.06, 2.75 | 0.38 |
| P-tau, pg/mL | 0.08 | -0.14, 0.31 | 0.46 |
| NfL, pg/mL | 2.04 | -3.62, 7.70 | 0.48 |

Note. Models were adjusted for age, sex, education, race/ethnicity, *APOE*- ε4 status, cognitive status, and GDS. $\beta$ Indicates the degree of change in outcomes per 1 unit increase in SCD. Bold font indicates *p*-value<0.05. \*FDR-adjusted *p*-value<0.05. Aβ , amyloid beta; APOE, apolipoprotein E; CSF, cerebrospinal fluid; FDR, false discovery rate; GDS, Geriatric Depression Scale; NfL, neurofilament light; p-tau, phosphorylated tau; SCD, subjective cognitive decline.

### *SCD x Sex* Interactions on CSF Biomarkers

SCD interacted with sex on Aβ42/40 (β=0.005, *p*=0.03) but not on any other CSF biomarkers (*p*-values>0.10). In stratified analyses, SCD was associated with Aβ42/40 in men (β=-0.006, *p*=0.0003), but not in women (*p*=0.48). These results were largely unchanged in sensitivity analyses excluding outliers but the overall interaction was attenuated after FDR correction (*p*=0.15). See **Table 3** for results and **Figure 2** for illustrations.

**Figure 2.**
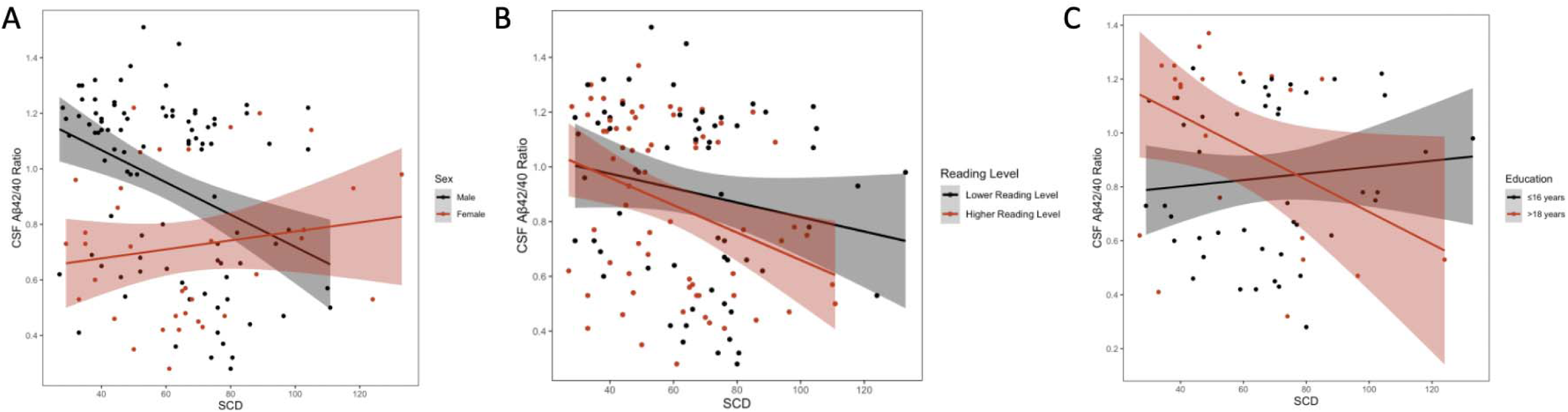
Subjective Cognitive Decline x Sex, Reading Level, and Education Interactions on Cerebrospinal Fluid Amyloid-β_42/40_ Ratio. Lines reflect CSF Aβ42/40 values corresponding to SCD levels. Shading reflects 95% confidence interval. (A) Associations between SCD and CSF Aβ42/40 ratio, stratified by sex; males β=-0.006, p=0.0003, females β=0.02, p=0.48. (B) Associations between SCD and CSF Aβ42/40 ratio, stratified by reading level; lower reading level β=-0.003, p=0.12, higher reading level β=-0.005, p=0.004. (C) Associations between SCD and CSF Aβ42/40 ratio, stratified by education; ≤16 years β=0.002, p=0.28, >18 years β=-0.006, p=0.23. Aβ, amyloid beta; SCD, subjective cognitive decline.

**Table 3.**
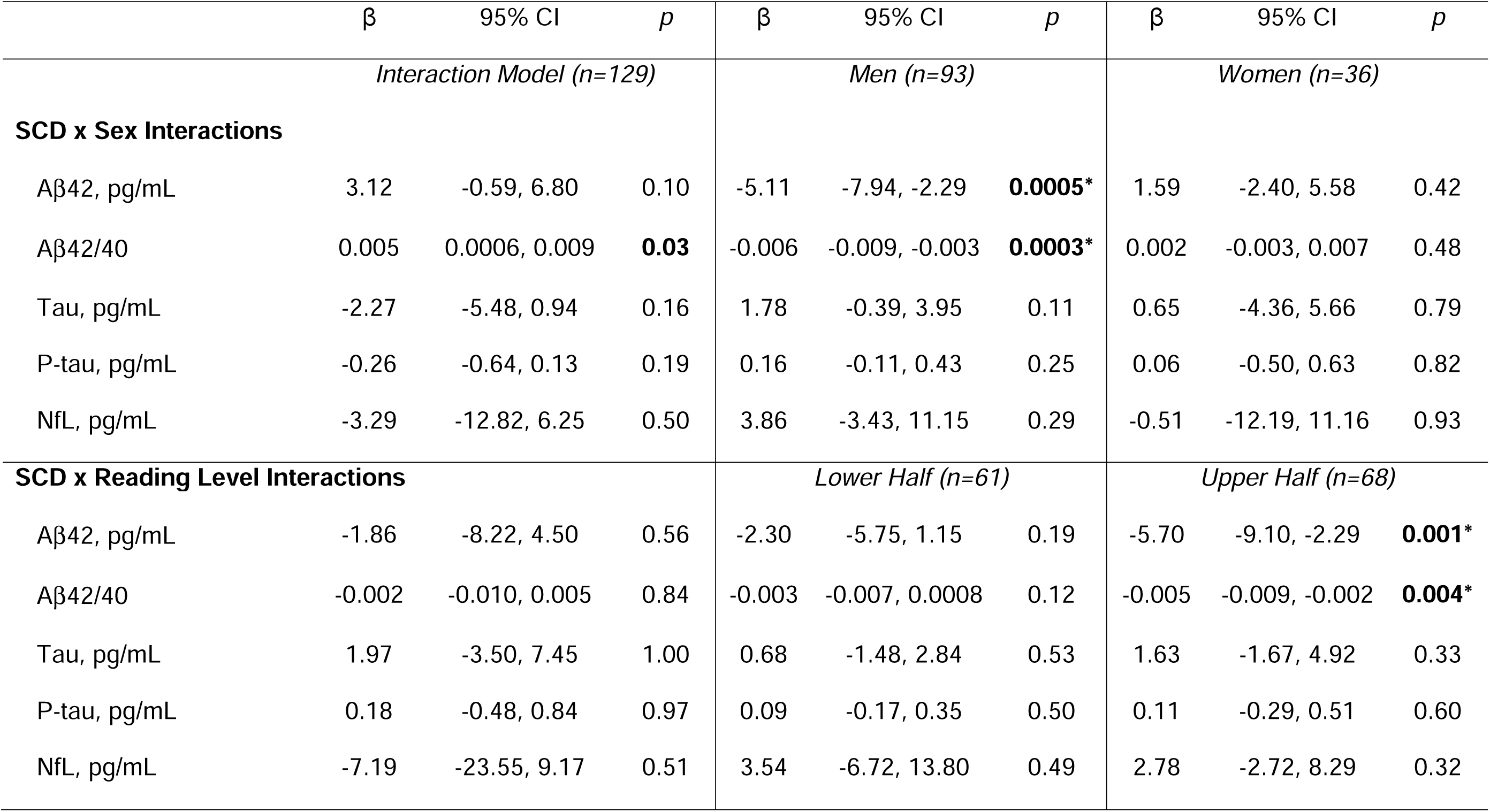
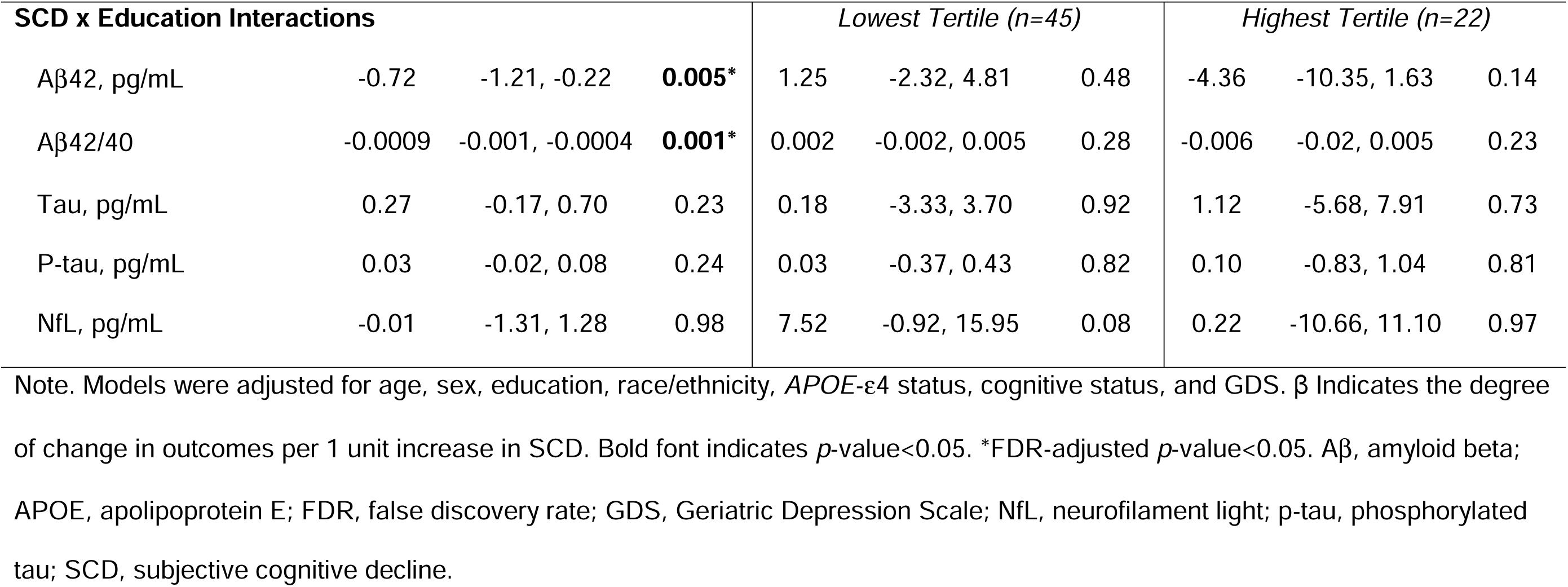
Interaction Models.

| | $\beta$ | 95% CI | $p$ | $\beta$ | 95% CI | $p$ | $\beta$ | 95% CI | $p$ |
| --- | --- | --- | --- | --- | --- | --- | --- | --- | --- |
| *Interaction Model (n=129)* |  |  |  | *Men (n=93)* |  |  | *Women (n=36)* |  |  |
| **SCD x Sex Interactions** |  |  |  |  |  |  |  |  |  |
| Aβ42, pg/mL | 3.12 | -0.59, 6.80 | 0.10 | -5.11 | -7.94, -2.29 | **0.0005\*** | 1.59 | -2.40, 5.58 | 0.42 |
| Aβ42/40 | 0.005 | 0.0006, 0.009 | **0.03** | -0.006 | -0.009, -0.003 | **0.0003\*** | 0.002 | -0.003, 0.007 | 0.48 |
| Tau, pg/mL | -2.27 | -5.48, 0.94 | 0.16 | 1.78 | -0.39, 3.95 | 0.11 | 0.65 | -4.36, 5.66 | 0.79 |
| P-tau, pg/mL | -0.26 | -0.64, 0.13 | 0.19 | 0.16 | -0.11, 0.43 | 0.25 | 0.06 | -0.50, 0.63 | 0.82 |
| NfL, pg/mL | -3.29 | -12.82, 6.25 | 0.50 | 3.86 | -3.43, 11.15 | 0.29 | -0.51 | -12.19, 11.16 | 0.93 |
| **SCD x Reading Level Interactions** |  |  |  | *Lower Half (n=61)* |  |  | *Upper Half (n=68)* |  |  |
| Aβ42, pg/mL | -1.86 | -8.22, 4.50 | 0.56 | -2.30 | -5.75, 1.15 | 0.19 | -5.70 | -9.10, -2.29 | **0.001\*** |
| Aβ42/40 | -0.002 | -0.010, 0.005 | 0.84 | -0.003 | -0.007, 0.0008 | 0.12 | -0.005 | -0.009, -0.002 | **0.004\*** |
| Tau, pg/mL | 1.97 | -3.50, 7.45 | 1.00 | 0.68 | -1.48, 2.84 | 0.53 | 1.63 | -1.67, 4.92 | 0.33 |
| P-tau, pg/mL | 0.18 | -0.48, 0.84 | 0.97 | 0.09 | -0.17, 0.35 | 0.50 | 0.11 | -0.29, 0.51 | 0.60 |
| NfL, pg/mL | -7.19 | -23.55, 9.17 | 0.51 | 3.54 | -6.72, 13.80 | 0.49 | 2.78 | -2.72, 8.29 | 0.32 |

| SCD x Education Interactions |  |  |  | Lowest Tertile (n=45) |  |  | Highest Tertile (n=22) |  |  |
| --- | --- | --- | --- | --- | --- | --- | --- | --- | --- |
| Aβ42, pg/mL | -0.72 | -1.21, -0.22 | **0.005\*** | 1.25 | -2.32, 4.81 | 0.48 | -4.36 | -10.35, 1.63 | 0.14 |
| Aβ42/40 | -0.0009 | -0.001, -0.0004 | **0.001\*** | 0.002 | -0.002, 0.005 | 0.28 | -0.006 | -0.02, 0.005 | 0.23 |
| Tau, pg/mL | 0.27 | -0.17, 0.70 | 0.23 | 0.18 | -3.33, 3.70 | 0.92 | 1.12 | -5.68, 7.91 | 0.73 |
| P-tau, pg/mL | 0.03 | -0.02, 0.08 | 0.24 | 0.03 | -0.37, 0.43 | 0.82 | 0.10 | -0.83, 1.04 | 0.81 |
| NfL, pg/mL | -0.01 | -1.31, 1.28 | 0.98 | 7.52 | -0.92, 15.95 | 0.08 | 0.22 | -10.66, 11.10 | 0.97 |

Note. Models were adjusted for age, sex, education, race/ethnicity, *APOE*- ε4 status, cognitive status, and GDS. $\beta$ Indicates the degree of change in outcomes per 1 unit increase in SCD. Bold font indicates $p$ -value<0.05. \*FDR-adjusted $p$ -value<0.05. Aβ , amyloid beta; *APOE*, apolipoprotein E; FDR, false discovery rate; GDS, Geriatric Depression Scale; NfL, neurofilament light; p-tau, phosphorylated tau; SCD, subjective cognitive decline.

### *SCD x Reading Level* Interactions on CSF Biomarkers

SCD did not interact with reading level on any CSF biomarkers (*p*-values>0.51). In stratified analyses, SCD was associated with Aβ42 (β=-5.70, *p*=0.001) and Aβ42/40 (β=-0.005, *p*=0.004) in the higher reading level group, but not in the lower reading level group (*p*-values>0.12). These results persisted after FDR correction and were largely unchanged in sensitivity analyses excluding outliers. See **Table 3** for results and **Figure 2** for illustrations.

### *SCD x Education* Interactions on CSF Biomarkers

SCD interacted with education on Aβ42 (β=-0.72, *p*=0.005) and Aβ42/40 (β=-0.0009, *p*=0.001), but not on any other CSF biomarkers (*p*-values>0.23). While the association between SCD and amyloid markers became stronger in individuals of higher education when education was considered as a continuous variable, there were no significant associations when results were stratified by educational tertile. These results persisted after FDR correction and were largely unchanged in sensitivity analyses excluding outliers. See **Table 3** for results and **Figure 2** for illustrations.

## Discussion

Among community-dwelling, nondemented older adults, higher levels of SCD were associated with decreased CSF levels of Aβ42 and a lower Aβ42/40 ratio. SCD was not associated with other CSF biomarkers of tauopathy or neurodegeneration. SCD interacted with sex on CSF Aβ42/40 such that associations were seen in men but not women, and SCD interacted with education such that associations were stronger in individuals of higher educational levels and educational quality. Taken together, these results highlight the relevance of SCD for screening nondemented older adults for AD pathological changes and suggest that demographic variables are important to consider when doing so.

Our findings add to the growing body of evidence supporting a link between SCD and amyloid deposition in non-demented older adults,^10,23–26^ and also demonstrate the clinical utility of a novel measure of SCD. The accumulation of cerebral amyloid is an early pathological event in AD, but its direct association with cognition has been questioned. Given the large number of cognitively normal individuals with evidence of amyloid pathology at autopsy,^27^ it has been assumed that amyloid is not directly associated with cognitive deficits in the absence of tau pathology. *In-vivo* studes using amyloid PET imaging have also demonstrated no significant differences between amyloid positive and negative cognitively unimpaired individuals on objective neuropsychological measures.^28^ While indvidual studies inconsistently observe cognitive impairment in amyloid positive preclinical AD patients, subtle cognitive deficits associated with amyloid pathology have been demonstrated in meta-analyses.^29,30^ It is possible that the subtle effects of amyloid on cognition are not consistently detectable by objective neuropsychological instruments. Measures of SCD are thought to be elevated at the earliest stages of AD^31^ and may be more attuned towards the subtle changes associated with amyloid accumulation. Beyond amyloid, we found that SCD is not associated with other biomarkers of tauopathy or neurodegeneration. These markers are typically associated with more significant cognitive decline and are closely linked to disease progression to MCI and dementia.^32^ As AD progresses, patients lose insight into their deficits, a phenomenon known as anosognosia. This loss of awareness limits the utility of self-reported cognitive changes and may explain why SCD was not associated with these biomarkers of more advanced disease.

We also found that associations between this novel SCD measure and CSF amyloid levels were varied across sex, with significant associations only being observed in men. This finding is surprising and contrary to past literature which suggests that SCD is more associated with clinical decline in women than men.^8^ There are a number of potential explanations for this finding. First, men generally are less likely to report or they tend to under-report the severity of cognitive symptoms.^33^ The current findings could suggest that when men endorse SCD, these reports are more accurately reflecting underlying pathology and amyloid deposition compared to women. Alternatively, these findings could represent a resilience to amyloidosis in women in the early stages of disease that is not present in men; however, this would be contrary to past work suggesting that women display greater clinical symptoms compared to men with similar levels of pathology.^34^ Similarly, as a group, men had higher levels of education than women in this cohort. However, this was statistically adjusted for and is not likely to fully explain this finding. Lastly, older women are more likely to experience multiple health problems than men,^35^ and multimorbidity is linked to worse cognitive functioning.^36^ It is possible, therefore, that SCD is more likely to be related to alternative etiologies other than AD in women than in men.

Further, we found that SCD was more strongly associated with amyloidosis in individuals with greater quantity and quality of education. These findings are consistent with past work which has shown that SCD is more associated with objective cognitive impairment^7^ and development of dementia^37–39^ in individuals of higher educational level and the association between SCD and amyloid deposition as seen on PET scans is stronger in more educated older adults.^40^ The current findings extend past work by suggesting the association between education with SCD and biomarker status exists regardless of educational metric (years of education vs. education quality). Given the high level of education attainment in this, and many other, cohorts, future research should examine the effect of educational quality in individuals with fewer years of education. Taken cumulatively, SCD in men and individuals with higher education attainment/quality appear more associated with amyloid accumulation.

This study has a number of strengths. As discussed above, this study utilized a novel SCD measure which has shown excellent psychometric properties, thereby increasing the ability to detect meaningful clinical changes. We utilized a well-characterized cohort and comprehensively assessed potential confounders. Further, we used core laboratories to analyze CSF using excellent quality control procedures with technicians blinded to clinical information. There were some limitations worth discussion as well. First, the cross-sectional nature of this study limits the ability to assess causality. Also, after performing an FDR correction, some findings were attenuated. This raises the possibility of false-positive findings and highlights the necessity of replicating these findings. Finally, this cohort was ethnically/racially homogenous and highly educated, thus limiting the generalizability of findings in diverse populations.

In sum, we demonstrated that this novel SCD measure is significantly associated with changes in CSF amyloid in nondemented older adults, with associations being stronger in men and in individuals with higher educational levels. These findings highlight the utility of self-report measures of SCD in older adults and provide some guidance as to which patient populations may be at greater risk of underlying AD pathology, thereby further advancing personalized medicine in AD and dementia care. Future research is needed to better understand the causes of sex and education-level differences in the association between SCD and AD to improve screening for early pathological changes across all patient populations.

## Data Availability

Due to participant consent limitations in data sharing, a subset of data is available for purposes of reproducing the results or procedures. These data, analytic methods, and study materials can be obtained by contacting the corresponding author.

https://www.vmacdata.org

